# A Genetics-guided Integrative Framework for Drug Repurposing: Identifying Anti-hypertensive Drug Telmisartan for Type 2 Diabetes

**DOI:** 10.1101/2025.03.22.25324223

**Authors:** Xue Zhong, Qiang Wei, Anshul Tiwari, Quan Wang, Yuting Tan, Rui Chen, Yan Yan, Nancy J Cox, Bingshan Li

## Abstract

Drug development is a long and costly process, and repurposing existing drugs for use toward a different disease or condition may serve as a cost-effective alternative. As drug targets with genetic support have a doubled success rate, genetics-informed drug repurposing holds promise in translating genetic findings into therapeutics. In this study, we developed a Genetics Informed Network-based Drug Repurposing via in silico Perturbation (GIN-DRIP) framework and applied the framework to repurpose drugs for type-2 diabetes (T2D). In GIN-DRIP for T2D, it integrates multi-level omics data to translate T2D GWAS signals into a genetics-informed network that simultaneously encodes gene importance scores and a directional effect (up/down) of risk genes for T2D; it then bases on the GIN to perform signature matching with drug perturbation experiments to identify drugs that can counteract the effect of T2D risk alleles. With this approach, we identified 3 high-confidence FDA-approved candidate drugs for T2D, and validated telmisartan, an anti-hypertensive drug, in our EHR data with over 3 million patients. We found that telmisartan users were associated with a reduced incidence of T2D compared to users of other anti-hypertensive drugs and non-users, supporting the therapeutic potential of telmisartan for T2D. Our framework can be applied to other diseases for translating GWAS findings to aid drug repurposing for complex diseases.

## INTRODUCTION

There is an increasing trend in the prevalence of complex chronic diseases in the United States and is expected to worsen in the next several decades ^1^. Currently, ∼50% of the US population has a chronic disease, with 86% of health care costs attributable to chronic disease ^1^. Diabetes, for example, is estimated to have a prevalence of 463 million people in 2019 and projected to be 700 million by 2045 ^2^. It is among the top 10 causes of death in adults with an estimate of four million deaths globally in 2017 ^3^. Type 2 diabetes (T2D) is the most prevalent form of the disease, accounting for more than 90% of the diabetes ^2^. T2D is very complex with highly heterogenous mechanisms, imposing challenges for disease management and treatment. Although there have been drugs for T2D, these drugs are often suboptimal given the complexity of the disease, demanding development of novel therapeutics. *De novo* drug development is however highly costive and often has high attrition rates. On the other hand, repurposing existing drugs to a different indication is a viable and cost-effective strategy ^4^. Successfully repurposed drugs include rituximab for rheumatoid arthritis ^5^ and sildenafil for erectile dysfunction ^6^. Although drug candidates for repurposing have met safety requirements, many have failed in clinical trials due to lack of efficacy ^4,7^. The challenge is mainly due to the lack of understanding of the disease mechanisms, as for complex diseases hundreds or even thousands of genes may be implicated in disease etiology, demanding novel approaches to dissect disease mechanisms for successful drug repurposing.

Drug targets with genetic support have been shown to increase the success rate by more than two folds ^8–10^. It is reported that two-thirds of the 50 drugs approved in 2021 have genetic support ^11^, showcasing the importance of genetics in drug development. Earlier computational methods utilize genetic support for drug repurposing ^12–14^ through identifying drugs whose targets overlap with disease risk genes. More recent approaches integrate genetic and transcriptomic data based on the principle that drugs capable of reversing the expression of disease risk genes are promising candidates ^13,14^. Nevertheless, the performance of these approaches relies on how accurate the disease risk genes can be uncovered, which remains a challenging task as the GWAS loci are generally located in the noncoding regions of the genome and do not automatically reveal the actual disease risk genes. Moreover, effective drugs may act not by targeting the disease risk genes themselves, but rather by targeting their interacting partners to correct and normalize the dysfunctional pathways (or subnetworks) implicated in the disease.

In this study we developed a computational framework, Genetics Informed Network-based Drug Repurposing via in silico Perturbation (GIN-DRIP), which integrates multiple levels of data from genetics, genomics, perturbation experiments and real-world electronic health records (EHRs) for drug repurposing. As a case study, we applied GIN-DRIP to repurpose FDA-approved drugs for T2D by utilizing T2D GWAS, TWAS and drug perturbation data, followed by validation studies of a top drug candidate in de-identified EHRs from 3 million patients. Our framework can be readily adapted to other diseases beyond T2D. As more GWAS loci are being discovered, GIN-DRIP has the potential to translate GWAS findings to guide effective drug repurposing.

## RESULTS

### GIN-DRIP: Genetics Informed Network-based Drug Repurposing via in silico Perturbation

We developed GIN-DRIP, a computational drug repurposing framework, and applied it to T2D in this study. The framework is summarized in **Fig. 1a**. A core component of the framework is to construct a T2D-specific genetics-informed gene network with nodes (genes) reflecting 1) the importance of genes in predisposition to T2D, and 2) the sign (+/-) of the effects of T2D risk alleles on the expression of the genes (up/down). Here we use node size to represent the genetic importance and node color for the up/down regulatory effect by T2D risk alleles (**Methods** and **Fig. 1**). Briefly, it starts with the identification of high-confidence risk genes (HRGs) for T2D GWAS loci, using iRIGS ^15^, a Bayesian integrative method that probabilistically predicts risk genes for each of the GWAS loci by integrating multi-omics data (**Fig. 1b**). It then uses the identified T2D HRGs as the seed genes and propagates their signals over the Gene Ontology (GO)-derived gene network to promote genes sharing pathways (and thus are “connected to”) with the seed genes in the network space, and assigns a score of importance to each gene; next, it infers the direction of regulatory effects (up/down) of T2D risk alleles on the risk genes from transcriptomic data of T2D relevant tissues (pancreas, adipose, skeletal muscle, liver) in the GTEx using TWAS ^16^ or PrediXcan ^17^. The result of the above steps is a signed genetics-informed T2D-specific network, which encodes genetic importance and up/down regulatory effects of T2D risk alleles on implicated risk genes (**Fig. 1c**).

**Fig. 1:**
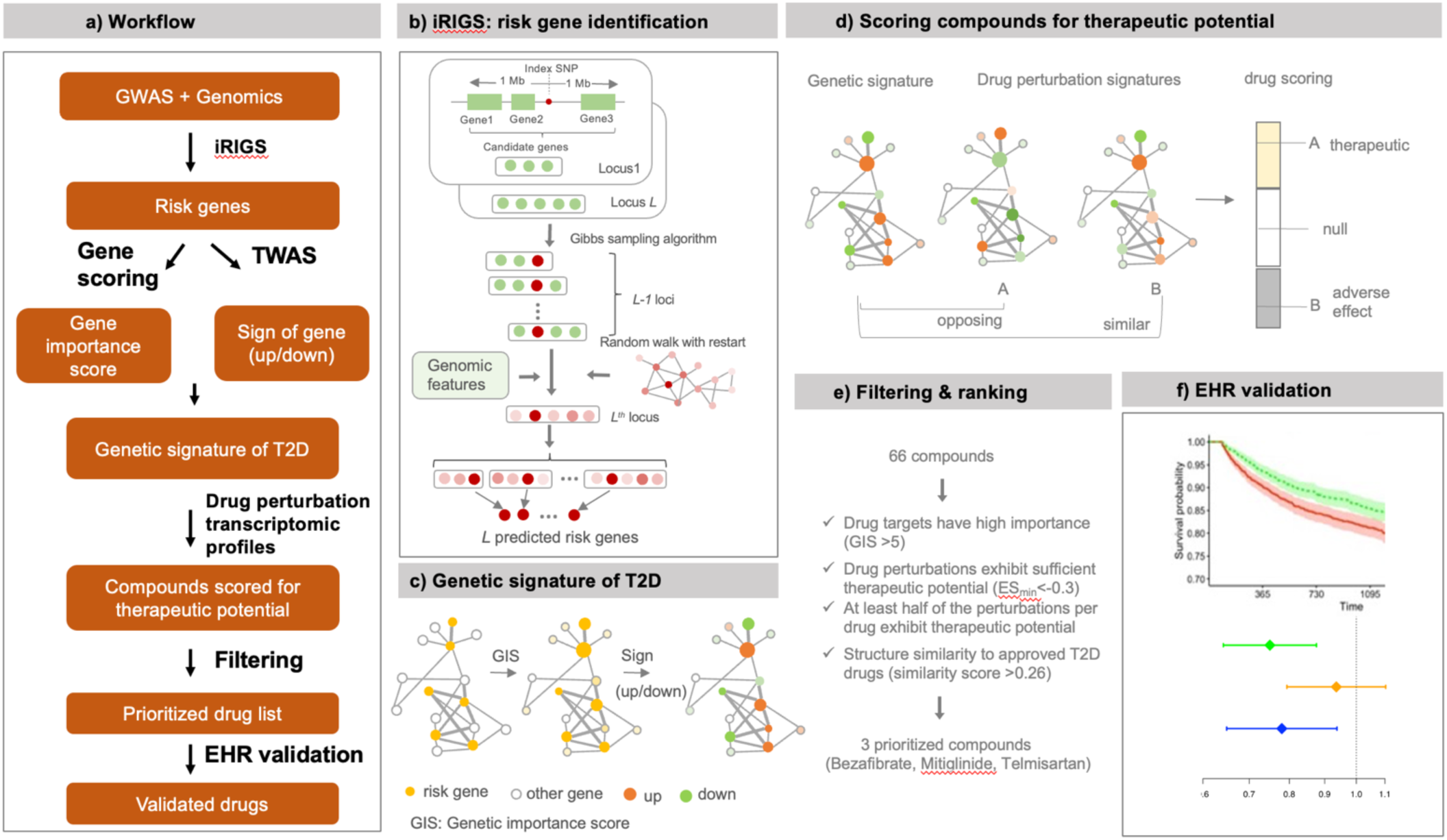
GIN-DRIP computational framework. **(a)** The overall workflow. **(b)** The iRIGS framework integrates GWAS loci, multi-omics data and gene networks to identify confidence risk genes of T2D. **(c)** T2D genetic signature via propagation of genetic risk genes through a GO-derived network. **(d)** Scoring of drugs by the drug’s potential of “reversing” T2D genetic signature using drug perturbation profiles from L1000 dataset. **(e)** Further prioritization of candidate drugs for validation. **(f)** Validation of high-confidence drug candidates in EHRs.

The next core task of our GIN-DRIP framework is to identify drugs that can reverse the sign of T2D signature genes as derived from the above T2D network (**Methods** and **Fig. 1**). Specifically, it extracts and calculates the transcriptional perturbation profiles of FDA-approved drugs available from the CMap ^18^ and L1000 ^19^ databases, aligns these drug perturbation profiles with the T2D genetic signature, and scores candidate drugs for the potential of “reversing” the T2D signature (**Fig. 1d**). After additional filtering and ranking, it narrows down to high-confidence candidate drugs and then validates the candidates in real-world patients EHR data (**Fig. 1e-f**). The details are described in the Methods section. In the following sections, we demonstrate the findings from each component of the GIN-DRIP framework with application to T2D.

### Bayesian integrative approach to infer risk genes underlying T2D GWAS loci

We collected 241 GWAS loci as reported in a large-scale T2D GWAS of 898,130 individuals of European decent ^20^. We curated multiple levels of genomic and epigenomic data relevant to T2D, including FANTOM5 ^21^, Capture Hi-C ^22^ and transcriptomic data in five T2D-relevant tissues (i.e. pancreas, liver, muscle and two adipose tissue types) from GTEx ^23^, as input features for iRIGS to probabilistically infer the underlying risk genes (see Methods for details of the model and algorithm). In total, we identified 235 unique HRGs (see **Suppl. Table S1** for the complete list of the genes and companying information). Among the list, we uncovered known T2D genes, e.g. *INS* ^24^*, IRS1* ^25^*, PIK3R1* ^26,27^, and *PIK3CA* ^28^ that are important in T2D pathophysiology. Gene-set enrichment analysis of the 235 HRGs in KEGG ^29,30^ revealed significant enrichment in pathways such as insulin resistance, insulin signaling and pancreatic secretion etc. (**Fig. 2a, Suppl. Table S2**), providing a solid foundation for genetics-informed drug repurposing for T2D at the genome-scale.

**Fig. 2:**
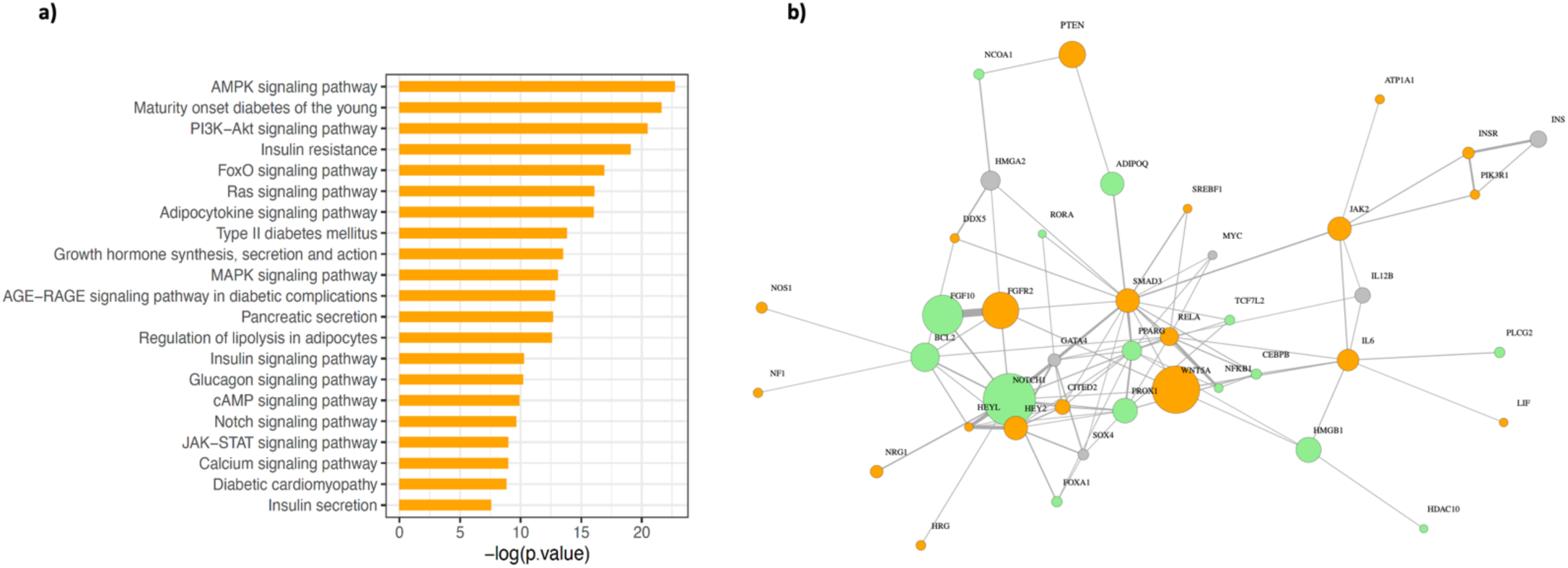
Pathway and network of T2D risk genes. **(a)** KEGG Pathways enriched in T2D risk genes. **(b)** A local signed genetic network centered at NOTCH1. Node color: orange, upregulated; green, downregulated; gray, direction unknown. Node size reflects the importance score of genes.

### Genome-wide prioritization of genes for T2D and construction of T2D-specific network

Beyond the genetic risk genes, their interacting partners in the same pathways also have potential to serve as drug targets. In addition, there are loci that remain uncovered as the current T2D GWAS loci explain only a small portion of the T2D heritability ^20^. Therefore, we leverage the identified HRGs and GO-derived network to prioritize genes at a genome-wide scale for T2D to facilitate drug repurposing. Genes that share more GO terms with HRGs are expected to receive higher genetic importance scores (GIS) (see **Methods** for details). The resulting genes ranked by the GIS are shown in **Suppl. Table S3**. The top-ranked gene is *NOTCH1*. The high score of this HRG is driven by the GO terms it belongs to that are highly enriched for other HRGs. We show that *NOTCH1* is a hub gene connecting to several HRGs with high GIS. These HRGs include known T2D genes such as *WNT5A* ^31^, *FGFR2* ^32^, *FGF10* ^33^, *PPARG* ^34^, *TCF7L2* ^35^ and *BCL2* ^36^. The T2D-specific subnetwork centering around *NOTCH1* is shown in **Fig. 2b**, in which the size of the node (gene) represents the GIS of the gene, and color represents the up (red) or down (green) regulatory effects by the T2D risk alleles. Notably, increased expression of *FGFR2* and *WNT5A* and decreased expression of *PPARG* are associated with T2D risk alleles. *NOTCH1* plays an important role in diabetes ^37,38^, and inhibition of *Dll4-Notch* signaling by anti-Dll4 has been shown to improve pancreatic islet function and insulin production ^39^. *PPARG* agonists act to boost insulin sensitivity and insulin action and have served as a component for several classes of T2D therapeutics including sulfonylurea, insulin secretagogue (meglitinide) and thiazolidinedione. These lines of evidence support the crucial role of this T2D subnetwork, demonstrating the effectiveness of our genetics-informed approach for scoring and identifying key genes and pathways involved in T2D.

### Identifying high-confidence candidate drugs with potential of repurposing for T2D

We used the compound-induced transcriptomic perturbation data from L1000 to identify FDA-approved drugs with the rationale that drugs capable of inducing gene expression changes in the opposite direction of T2D gene signature have therapeutic potential for treating T2D (**Fig. 3a**, also see **Methods**). The L1000 platform contains 978 landmark genes and 11,350 inferred genes with 720,216 perturbation combinations (perturbagen type, duration, concentration etc.), including FDA-approved T2D drugs ^19^. For each FDA-approved drug assayed, we calculated a repurposing score, which quantifies the drug’s potential of reversing the T2D risk gene effects (**Methods**). To evaluate the effectiveness of this approach, we used FDA-approved T2D drugs as positive controls to test the rationale. We compared the repurposing scores for the T2D drugs using genes with high GIS (i.e. GIS>5, 54 genes) vs. low GIS (GIS in the lowest 25 percentile, 3592 genes), and observed that the repurposing scores of the T2D drugs are significantly greater for genes with GIS>5 than for the background (*P* = 0.034). We also evaluated it using GIS>6 and GIS>4 as the cutoff to define the T2D signature and observed similar results (*P* = 0.015 and 0.0056 respectively).

**Fig. 3:**
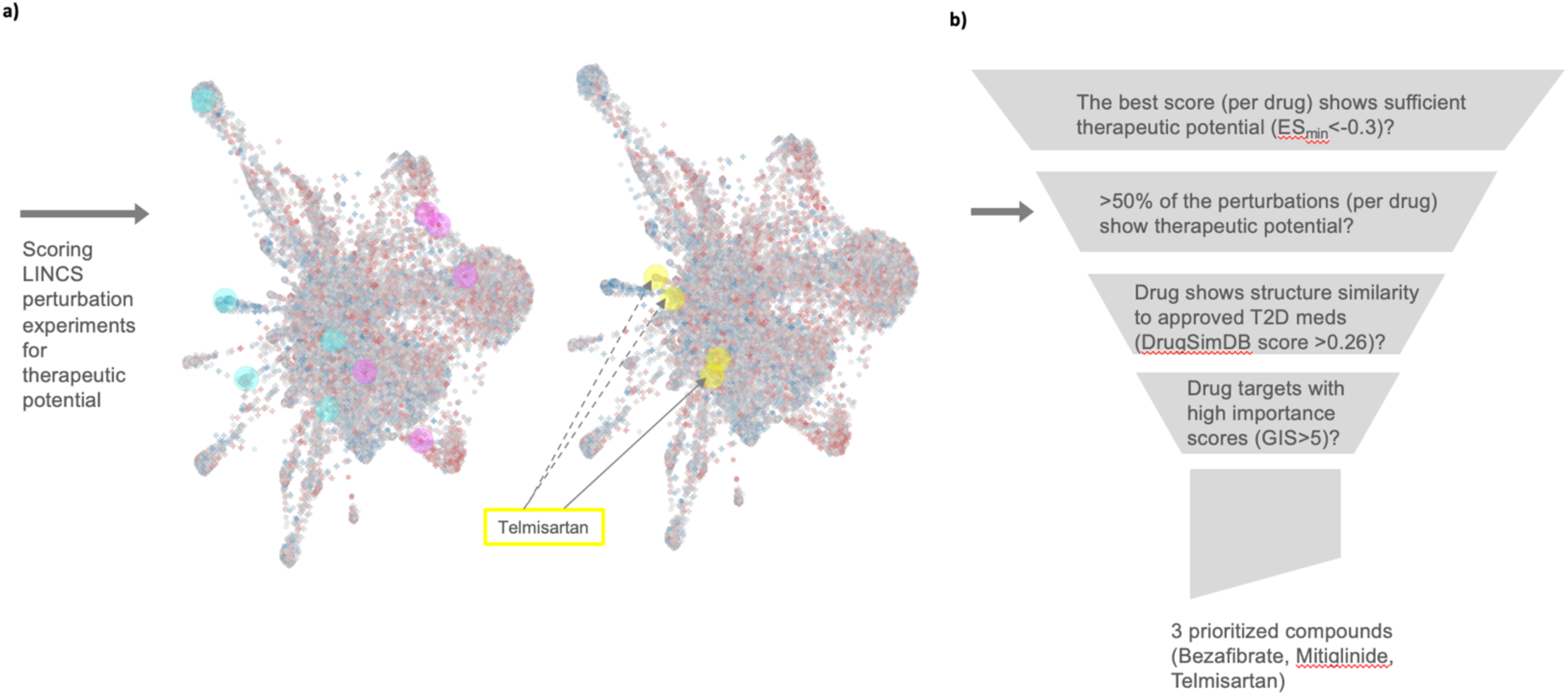
Rank LINCS L1000 small molecules that integrate data of bioactivity and chemical structure. **(a)** Rank L1000 small molecules based on the similarity between the compound transcriptional response signatures and T2D genetic signature. Each node represents a transcriptional response signature; color purple indicates compounds showing similarity to the disease signature while color blue indicates a reversal effect of the disease signature. In general, a compound can present signatures varying across experimental conditions (cell-line, dose, duration). For example, telmisartan shows signatures, highlighted in yellow, of reversal effect (marked by the arrow with a solid line) in some conditions and null effect in other conditions (arrow with a dashed line). **(b)** Further prioritization based on the maximum and consistency of the reversal effect of the small molecules across various experimental conditions, their structure similarity to existing T2D medicines (DrugSimDB score >0.26) and importance of the drug targets in T2D. Telmisartan emerges as a top candidate after the comprehensive ranking method.

Based on the proven rationale using T2D drugs, we calculated repurposing scores of FDA-approved drugs by aligning drugs’ perturbation signature against T2D signature (defined as GIS>5) and identified top scoring drugs as candidate drugs repurposable for T2D. In total, we identified 66 drugs as candidates for T2D, spanning multiple categories of drugs (e.g. anti-hypertensive, lipid-lowering, anti-inflammatory, weight-loss) (**Suppl. Table S4)**. A literature search of these drugs showed multiple lines of support for some of the drugs. For instance, Ivacaftor, a medication belonging to cystic fibrosis transmembrane conductance regulator (CFTR) potentiators, improves insulin secretion ^40^ or reduces the glucose level ^41,42^ in patients with cystic fibrosis; Bezafibrate, a lipid-lowering drug, reduces blood glucose in T2D patients ^43–45^, reduces insulin resistance ^46^, decreases the development and delays the onset of type 2 diabetes ^47,48^; Telmisartan, an angiotensin II receptor antagonist, is an anti-hypertensive drug and has been reported to have beneficial effects on T2D, including improving insulin resistance through regulating glucose and lipid metabolism ^49–51^, improving hyperglycemia-induced cardiac fibrosis ^52,53^, preventing diabetic cardiomyopathy ^54,55^, preserving cardiovascular and renal function in patients with T2D ^56^. We further filtered the candidate drug list by three criteria: i) the protein sequence similarity between a candidate drug and FDA-approved T2D drugs is no worse than the within-T2D drug similarity on average, as calculated from DrugSimDB ^57^; ii) the candidate drug should exhibit sufficient therapeutic potential as quantified by its best (minimum) repurposing scores across experimental conditions; and iii) the GIS of the drug targets. We finally identified 3 drugs (bezafibrate, mitiglinide, and telmisartan) as high-confidence candidates (**Fig. 3b; Methods**). Of a particular note, mitiglinide has been approved to treat T2D in Japan but not yet by the FDA.

### Effect of telmisartan on T2D incidence in VUMC EHRs

Telmisartan is widely used to treat hypertension. Given the relatively large number of telmisartan users, we set out to evaluate the effect of telmisartan on the incidence of T2D in Vanderbilt University Medical Center (VUMC) de-identified EHR database containing medical records from ∼3 million patients. We defined telmisartan-exposed patients as those with at least one year of documented telmisartan use in their EHRs. To alleviate confounding by indication, we applied an active comparator design ^58^, by selecting users of other anti-hypertensive drugs to construct comparators for telmisartan. The selected anti-hypertensive drugs differ from telmisartan in their mechanisms of action ^59^, including clonidine (an alpha-agonist anti-hypertensive drug) and diltiazem (a calcium channel blocker anti-hypertensive drug), while telmisartan belongs to the angiotensin II receptor blocker category. For each cohort (telmisartan, clonidine and diltiazem), we only included individuals who were on a single arm of the hypertensive drug. We also constructed a pooled non-user (of telmisartan) cohort that consists of those who do not use telmisartan (but can use other anti-hypertensive drugs). Given that hypertension is a prevalent condition ^60^ and is affected by age ^61^ and obesity ^62–64^, all cohorts excluded individuals with age<20 years and/or BMI<18 at baseline before the drug initiation.

We further used propensity score (PS) matching on gender, age (birth year) and comorbid conditions relevant to T2D including obesity, hyperlipidemia, and coronary artery disease (CAD) to minimize potential confounders between cohorts (**Methods**). Consequently, we obtained cohorts of telmisartan users (n=2257), users of clonidine (n=4511), diltiazem (n=4458) and pooled non-users (n=9028). **Table 1** summarizes the patient characteristics for each cohort. At baseline, all cohorts constructed are similar in age (60 yr **±** 13), gender (49% male) and prevalence of comorbid conditions (∼65% hyperlipidemia, ∼34% CAD and ∼87% hypertension).

**Table 1:** Characteristics of cohorts for telmisartan validation in VUMC EHRs.

|  | **Telmisartan**<br>(N=2257) | **Clonidine**<br>(N=4512) | **Diltiazem**<br>(N=4489) | **Non-user**<br>(N=9028) |
| --- | --- | --- | --- | --- |
| **Baseline characteristics** |  |  |  |  |
| Age (Mean, SD) | 60.0<br>(13.0) | 59.9<br>(12.9) | 60.4<br>(12.8) | 59.9<br>(13.0) |
| Sex (Male, n, %) | 1097<br>(48.6%) | 2172<br>(48.1%) | 2140<br>(47.7%) | 4388<br>(48.6%) |
| BMI (Mean, SD) | 29.9<br>(4.96) | 29.7<br>(4.89) | 29.9<br>(4.90) | 29.9<br>(4.95) |
| Hyperlipidemia | 1459<br>(64.6%) | 2920<br>(64.7%) | 2884<br>(64.2%) | 5836<br>(64.6%) |
| Coronary artery Disease | 755<br>(33.5%) | 1508<br>(33.4%) | 1515<br>(33.7%) | 3020<br>(33.5%) |
| Hypertension | 1873<br>(83.0%) | 3746<br>(83.0%) | 3730<br>(83.1%) | 7492<br>(83.0%) |
| **Outcome** |  |  |  |  |
| Newly diagnosed T2D | 651<br>(28.8%) | 1528<br>(33.9%) | 1252<br>(27.9%) | 2425<br>(26.9%) |

We estimated Kaplan-Meier curves for each comparison and conducted propensity score stratified Cox regression analysis. As shown in **Fig. 4**, after 5 years of follow-up, the telmisartan usage was significantly associated with reduced risk of T2D compared to the non-user group (**Fig. 4** top left panel; hazard ratio [HR]=0.78, 95% confidence interval [CI]=0.71–0.86, *P*=2×10^-7^). In the active comparator analysis, telmisartan usage was significantly associated with reduced risk of T2D compared to clonidine (**Fig. 4** top right panel; HR=0.74, 95% CI=0.67–0.83, *P*=2.5×10^-8^) but not diltiazem (**Fig. 4** bottom left panel; HR=0.96, 95% CI=0.86-1.07, *P*=0.47). The results were similar in sensitivity analyses stratified by gender age groups: no difference was detected between males and females (**Suppl. Fig. 2**), or between older (age>55 years old) and younger patients (age 20-55 years old) (**Suppl. Fig. 3**).

**Fig 4.**
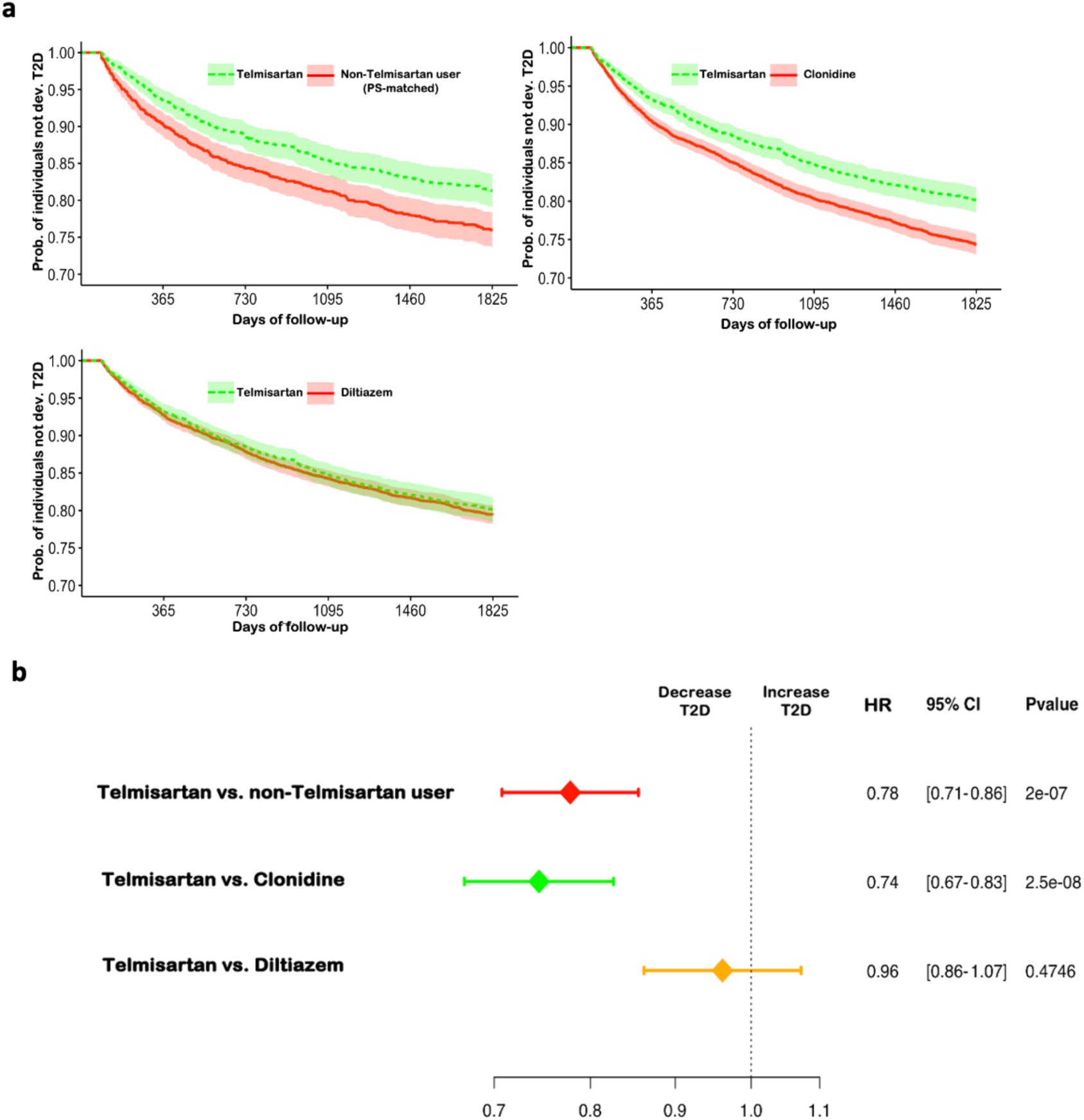
EHR analysis of telmisartan on T2D. **(a)** Survival analyses reveal that the usage of telmisartan is associated with reduced incidence of T2D in VUMC EHRs. **(b)** Hazard ratios (HR) and 95% confidence interval (CI) across three comparison studies. Telmisartan, clonidine and diltiazem are anti-hypertensive drugs with different mechanisms of action.

## DISCUSSION

Massive advances in GWAS provide unprecedented opportunities for understanding disease mechanisms and to aid drug development. Nevertheless, it remains challenging given the increasingly recognized complexity of disease. GWAS do not directly reveal pathophysiological information or risk genes for a disease ^12^. In this study, we developed GIN-DRIP, a novel computational framework that integrates multi-levels of data to translate GWAS findings and to aid the search of therapeutics. As a case study, we applied GIN-DRIP to T2D to identify and validate drugs potentially repurposable for T2D. T2D is a complex disease, and hundreds of genes are involved ^20^. Although many drugs with different mechanisms of action are available to treat T2D, the management of the efficacy, side effects and complications is difficult to achieve ^65,66^. It is proposed that reducing HbA1c is unlikely to be optimal treatment, and drugs or multiple drugs that target the multifaceted pathogenic abnormalities may be needed to treat the disease ^67^. Our genetics-informed systemic approach in GIN-DRIP prioritized genetically important risk genes for T2D on the genome scale and identified 66 initial candidates and prioritized three high-confidence candidate drugs for T2D. We selected telmisartan, an anti-hypertensive drug, for further validation in VUMC de-identified EHR data, and observed that telmisartan users showed significantly reduced incidences of T2D compared to hypertensive patients taking other anti-hypertensive drugs (clonidine).

Here we further dissect the value of GWAS for drug development, the challenges, and the added value of our work to the field. Rare protective loss of function (LoF) mutations have successfully informed drug development, e.g. approval of PCSK9 inhibitors alirocumab (Praluent) and evolocumab (Repatha) drugs that mimic the beneficial effect of *PCSK9* LoF mutations on lowering low-density lipoprotein (LDL) ^68^. GWAS of common variants can guide therapeutics in the same principle, albeit indirectly, as the GWAS loci are often located in the noncoding region and affect disease risk through regulation of the underlying (often unknown) risk genes. Promising drugs are expected to mimic the effect of protective alleles (or reverse the effect of risk alleles) to achieve therapeutic effects ^12,69,70^. Linking GWAS loci to the actual risk genes under regulation has proven challenging. TWAS ^16^ or PrediXcan ^17^ approaches have been used to identify candidate risk genes but they are often confounded by LD and co-regulation of genes within GWAS loci ^71,72^, bearing challenges for drug repurposing ^12^. Our approach integrates multiple levels of functional genomics data to robustly infer risk genes, and for T2D we demonstrated the accuracy of the inferred risk genes from multiple angles. Genetic risk genes may not always be the ideal drug targets, and interacting partners in the same pathways are often better drug targets. For example, one drug approved in 2021 is the *IFNAR1* antagonist anifrolumab for the treatment of systemic lupus erythematosus (SLE). While the authors did not find direct genetic support for *IFNAR1*, they instead found that missense variants in *TYK2* - a kinase that physically interacts with *IFNAR1* - has been previously associated with SLE ^73^, highlighting the potential utility of pathway and network for boosting the use of human genetics for drug development and repurposing ^11^. We took advantage of our inferred risk genes and GO-derived networks to construct a signed genetics-informed T2D network, prioritizing genetically important genes and subnetworks on the genome-scale for effective drug targeting, directly or indirectly. We further augmented the T2D genetics-informed network by providing the effects (up/down) of regulation of risk genes in T2D. Here, our use of TWAS/PrediXcan is different from previous approaches ^13,14^, in that our goal is not to identify risk genes but rather to evaluate the direction of risk genes on disease, avoiding the confounding encountered by traditional TWAS/PrediXcan. Our genetics-informed disease networks encode information about genetic risk genes, including the importance scores and the sign effects with respect to disease risk, which are fundamentally distinct from traditional network-based drug repurposing approaches ^74,75^. Since the genetic risk genes (direct or indirect) in the network are equipped with the sign of disease risk genes, the signature matching with drug perturbation experiments is able to distinguish therapeutic vs. adverse effects, addressing the challenges encountered in GWAS-based drug repurposing ^76^.

T2D is a highly heterogenous disease, and to date over 700 GWAS loci have been identified ^77^. For some patients, it is likely that multiple drugs targeting distinct components of the disease etiology may be needed for effective treatment. It is known that disease-risk genes function in distinct contexts at the levels of organs, tissues, cell types or subtypes. Single-cell RNA-seq (scRNAseq) and ATAC-seq (scATACseq) across various tissues are being generated for T2D, revealing islet cell types implicated in T2D ^78^ as well as beta cell subtypes and cell state shifting during T2D progression ^79^. As drugs eventually act on individual cells in patients, a future augment of our approach is to incorporate multi-omics data at single-cell resolution to build cell type or subtype-level disease-specific networks to facilitate cellular context-aware drug repurposing.

We used VUMC’s de-identified EHR database with over 3 million patients to validate the antihypertensive drug telmisartan as a candidate drug repurposing for T2D. Real-world clinical information in particular the longitudinal data accumulated over years in EHRs can serve as a valuable tool for repurposing existing drugs and discovery of adverse effects ^80,81^. Meanwhile, challenges of using EHR data for this purpose are well recognized for the need of effective control for biases and confounders ^82^. For drug repurposing in EHRs, drug use is often associated with patients’ health conditions, and as a result, baseline characteristics of patients taking a drug often differ systematically from those of untreated subjects or those taking different drugs. Propensity-score matching methods have been widely used to reduce confounding in observational studies ^83^. PS matching alone is unlikely to effectively control for all confounding factors, e.g. confounding by indication. To minimize potential confounders, we implemented in this study several strategies, including PS matching, matching by indication and the active comparator design. We validated telmisartan in VUMC’s EHR data using all of these strategies, strengthening the evidence supporting telmisartan as a potential T2D drug. Although these findings are promising, further validation in prospective clinical trials will be essential to fully establish telmisartan’s potential efficacy in T2D.

This study has several limitations. First, the T2D drug repurposing candidates we interrogated are limited to the FDA-approved small molecules tested in the LINCS database. Although a benefit of focusing on FDA-approved drugs is the known safety profiles of these compounds, the consequent search space is admittedly limited. Moreover, the cell types where the compounds were tested in LINCS have been primarily in cancer cell lines, which may not accurately recapitulate the gene expression patterns in T2D-relevant cell types. Future studies that expand beyond FDA-approved agents would require a pipeline of screening for adverse effects of novel compounds.

## METHODS

### Identification of 235 HRGs for T2D GWAS loci using integrative genomic approaches

We collected 241 GWAS loci for T2D with a sample size of 898,130 individuals of European decent ^20^. We used iRIGS, a Bayesian framework that integrates multiple levels of genomic data and a gene-gene network, to probabilistically infer risk genes for each of the GWAS loci. Briefly, iRIGS is to calculate the posterior probability (PP) of each candidate gene in the window centered around the GWAS index SNP and select the gene with the maximum PP as the risk gene for that locus. To calculate PP, iRIGS integrates two levels of information: 1) gene-gene networks and 2) gene-level genomics data, with the rationale that i) risk genes tend to converge to core pathways, and ii) risk genes have genomic features distinct from non-risk genes. We used a gene-gene network derived from Gene Ontology (GO), and gene-level features including FANTOM5, Capture Hi-C and distance to the index SNPs by rank as used in the original iRIGS ^15^, as well as from T2D-relevant tissues (adipose, liver, muscle and pancreas) from GTEx ^23^. For GTEx data, we calculated each gene’s specificity in each tissue and used the gene’s tissue specificity as the T2D-specific features. Accordingly, iRIGS uses a Gibbs sampler to calculate a one-dimensional conditional distribution to speed up the computation – for details see ^15^. For each GWAS locus of T2D, we defined a 2Mb window centered around the GWAS index SNP and extracted all protein-coding genes within the window as the candidate genes of that locus. When the locus window harbors more than 20 genes, we limited the candidates to a maximum of 20 nearest genes (from the index SNP). In total, we obtained 3198 candidate genes from the T2D GWAS loci.

### Construction of a signed genetics-informed T2D network

#### Defining importance scores for all genes based on HRGs and Gene Ontology

First, we collected genes in each of the GO term annotations (version 2021-05-01) and calculated the number of T2D HRGs in each of the GO terms; we use *h_i_* to denote the number of HRGs in the *i*^th^ GO term, *n_i_* to denote the total number of genes in the *i*^th^ GO term, and *T* to denote the total number of GO terms. We define the importance score of the *i*^th^ GO term as 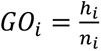, i.e. the fraction of HRGs in the GO term, similar to the approach used in NETBAG ^84^. Next, for each gene, we identified the GO terms containing that gene, and defined the gene’s genetic important score (GIS) as 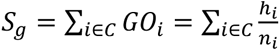, where *C* is the set of GO terms containing the gene. For the connection (edge) between genes, we summed all the shared GO term scores between genes. Of a pair of genes without sharing any GO term, their edge is zero (i.e. unconnected). Thus, we can construct a genetics-informed T2D-specific network (not signed yet), in which the node (gene) size represents the importance of the gene for T2D, and the edge between a pair of genes represents how tight the gene pair are connected.

#### Defining the sign (up/down) effects of risk genes on T2D

In the above T2D network, the node size encodes the importance of the gene to T2D but does not encode the sign effects of the risk gene, i.e. it is unknown whether the inhibition or activation of the gene predisposes T2D risk. Here we used the TWAS ^16^ approach to assess the association between gene expression and T2D risk alleles in five T2D-relevant tissues (pancreas, adipose subcutaneous, adipose visceral omentum, skeletal muscle, and liver), to resolve this issue. If risk alleles increase the expression of the gene, the activation of the gene predisposes T2D risk, and vice versa. In TWAS, we built prediction models for all genes in each of the five tissues, using Elastic Net. Specifically, we used the TPM as the quantification of gene expression, and SNPs in the +/- 500kbp windows of the genes as the predictors. As our primary goal is to assess the sign effects of the risk alleles on gene expression, we coded the T2D risk allele as ‘1’ and otherwise ‘0’ based on the T2D GWAS summary statistics. We kept for downstream analyses those gene expression models showing correlation of r>0.1 between predicted and measured gene expressions. We then performed association analysis using the predicted models with T2D GWAS summary statistics ^20^ and obtained association *P-values* and *Z-scores.* For the five T2D relevant tissues, if one or more tissues have *P*-value <0.05, we averaged the TWAS Z-scores of these tissues with *P*-value <0.05; if none of the tissues’ *P*-value <0.05, we averaged the TWAS Z-scores of all five tissues. If the summed TWAS Z-score is negative, the gene is considered being down-regulated (-1) in T2D, and conversely, being up-regulated (+1) in T2D.

### Identification of T2D candidate drugs using the signed genetics-informed T2D network and drug perturbation experiments

We used the LINCS L1000 project data to evaluate a drug’s potential to act on T2D genetically important genes to reverse the effects of T2D risk genes. The L1000 generated transcriptomics data of 978 landmark genes and 11,350 inferred genes in a variety of cell lines with a total of 720,216 perturbation experiments ^19^, including FDA T2D drugs such as Sulfonylurea and Meglitinide. Using different cutoffs of GIS, we obtained different gene sets to build T2D genetic signatures along with the sign of the genes as determined by TWAS. For a given T2D signature, we calculated a weighted Kolmogorov-Smirnov enrichment score (ES) for the assayed compounds as described in the L1000 paper ^19^. A negative value of ES indicates therapeutic potential while a positive ES indicates exacerbation potential. Multiple ES values were obtained for a compound with multiple perturbation experiments varying by duration, doses and cell lines, and we used the best ES (i.e., minimum ES or ES_min_) as the representative score for that compound. The ES score quantifies the enrichment of the T2D important genes in each perturbation-induced expression response profile. It calculates enrichment scores for the input gene set against each perturbation expression profile. It ranks genes and determine the proportion of the in-signature genes that rank higher than expected. The ES scores were visualized by L1000FWD ^85^.

We used genes whose GISs are higher than 5 (**Suppl. Table S3**) to identify candidate drugs. We performed TWAS ^16^ to evaluate the T2D risk alleles’ effect on expression of these 54 genes, i.e., up- or down-regulation of gene expression, and generated a total of 28 up-regulated and 26 down-regulated genes as a query gene list. To determine the threshold of ES_min_, we collected 27 FDA-approved T2D drugs in L1000 and calculated the ES_min_ of them. We found that the ES_min_ of all approved drugs are less than -0.3, therefore we used ES_min_< - 0.3 to select high-confidence candidate T2D drugs. We observed that the approved T2D drugs scored better against T2D signatures built by genes with high GIS (e.g. GIS>5, 54 genes) than with low GIS (GIS in the lowest 25 percentile, 3592 genes) (*P*=0.034). We also evaluated it using GIS>6 and GIS>4 as the cutoff and observed similar results (*P*=0.015 and 0.0056 respectively). Thus, we calculated repurposing scores of the FDA-approved drugs by aligning their perturbation signatures against the T2D signature defined as GIS>5 and identified top-scoring drugs.

In addition, we leveraged drug-drug similarity ^57^ to further narrow down the list. We utilized the database DrugSimDB ^57^ to calculate protein sequence similarities of drug targets between candidate drugs and FDA-approved T2D drugs. For each candidate drug, we took the mean value of similarities with the 27 approved drugs as its drug similarity measurement score. To determine the threshold of drug similarity, we evaluated the drug similarity scores of 27 approved drugs within themselves and found the drug similarity scores of all approved drugs are higher than 0.26. We therefore used a drug similarity score of 0.26 as the cutoff to identify high-confidence candidate drugs. Finally, we used the mean GIS of the drug targets to select drug candidates that target genetically important genes. Specifically, we used GIS greater than 5 as the cutoff for selecting high-confidence drug candidates. Combining GIS, ES_min_ and drug similarity score scores together, we finally obtained high-confidence candidate drugs by requiring GIS>5, ES_min_ <-0.3, and drug similarity score>0.26.

### Clinical validation: EHR database and cohort construction

We carried out a validation study of telmisartan in VUMC-SD, a hospital-based de-identified patient EHR database that has accumulated over decades for billing codes (of disease diagnosis and medical procedures), lab results and medication use ^86^. To avoid confounding by indication, we applied an active comparator design ^58^ and selected users of other anti-hypertensive drugs to construct comparators for telmisartan, including clonidine and diltiazem. For each cohort (telmisartan, clonidine and diltiazem), we only included those who were on a single hypertensive drug. We also constructed a pooled non-user (of telmisartan) cohort that consists of patients who do not use telmisartan (but can use other anti-hypertensive drugs). For all cohorts we excluded individuals with age<20 years and/or BMI <18 at baseline before the drug initiation. We restricted all our analyses to patients with hypertension.

Next, we define a drug episode for an individual exposed to a drug of interest. A drug episode is defined as the time interval from drug initiation (i.e., the first prescription date of that drug in the patient’s EHRs) to drug discontinuation (**Suppl. Fig. 1**). Drug discontinuation is defined if the time between two consecutive dates of prescriptions is greater than a pre-defined time gap ^87^. Here, we chose 365 days as the allowable gap and the first drug episode as the drug episode per user.

The primary outcome is the development of T2D during the observation window (i.e. the drug episode). The censoring time is the first diagnosis code of T2D or the end of the drug episode, whichever comes first. To further reduce confounding factors, we matched between cohorts the pre-treatment covariates that may affect the outcome (i.e., development of T2D), including age, gender, BMI, hyperlipidemia and coronary artery disease (CAD). This was done by propensity score (PS) matching ^83^ of the covariates at baseline (before the initiation of the drug) with a ratio up to 1:4 between telmisartan and other drug users. International Classification of Disease (ICD) codes used to identify the comorbid conditions and diagnosis of T2D ^88^ were listed in **Suppl. Table S5**.

### Statistical analysis

A Kaplan-Meier estimator method was used to estimate the time to T2D curves. Propensity score survival analyses were carried out to investigate the risk of T2D in users of telmisartan relative to clonidine, diltiazem and pooled non-users. Logistic regression was used to estimate the propensity scores for taking telmisartan, with covariates including age, sex, hyperlipidemia and CAD. Propensity score Cox proportional hazards models were used for statistical inference of the hazard ratios (HR) of developing T2D between telmisartan users and other cohorts.

## Data Availability

All data produced in the present work are contained in the manuscript.

http://diagram-consortium.org

https://predictdb.org

https://clue.io/data

https://go.drugbank.com

## Data availability

All data generated or analyzed during this study are included in this published article and its Supplementary Information. Access to VUMC EHR’s database requires institutional approval and compliance with a data use agreement. The following links provide the publicly available datasets we used. The T2D GWAS summary statistics are available at http://diagram-consortium.org.The genotype-based gene expression imputation (TWAS) models can be downloaded from https://predictdb.org

The LINCS drug perturbation data can be downloaded at: https://clue.io/data/CMap2020#LINCS2020

Annotation of drugs were collected from DrugBank: https://go.drugbank/com/

## Code availability

The code and companion data used are in https://github.com/cgg-L/GIN-DRIP

## Acknowledgement

The study is supported by the National Institute on Aging of National Institutes of Health under award numbers R01AG069900, R01AG089717 and R01AG065611.

## Author contributions

B Li and X Zhong conceived and supervised the study. Q Wei collected and downloaded data. Q Wei, Q Wang and X Zhong performed the T2D risk gene discovery and drug analysis; A Tiwari performed the EHR validation analysis; X Zhong, Q Wei, A Tiwari and Q Wang prepared the Figures and Tables. B Li, X Zhong, Q Wei, A Tiwari and Q Wang wrote the manuscript. All authors participated in the interpretation of the results, read and/or edited the final manuscript. All authors approved the final version of the manuscript and consent for publication.

