## Supplementary_figure_S1-S3 for "A Genetics-guided Integrative Framework for Drug Repurposing: Identifying Anti-hypertensive Drug Telmisartan for Type 2 Diabetes"

**Supplementary Figures**

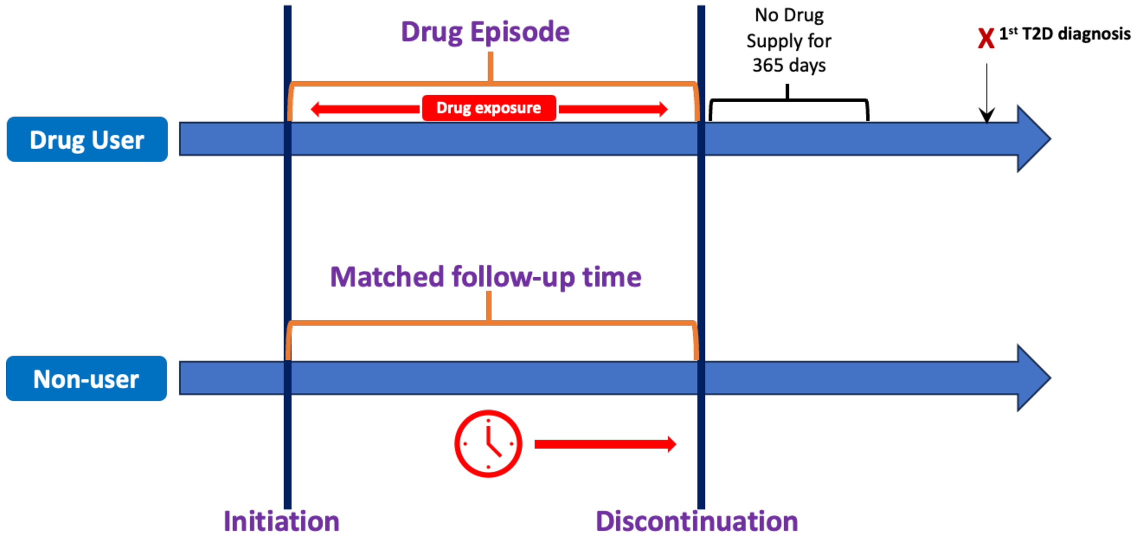


**Supplementary Fig. 1: Definition of a drug episode**. A drug episode is defined as the interval of time between the initiation of the drug and its discontinuance. The first day of drug supply is considered drug initiation (i.e., 1st prescription date). The last day of drug supply is defined as drug discontinuation (i.e., last prescription date + days of supply) and without medication for the following 365 days.

**a)**

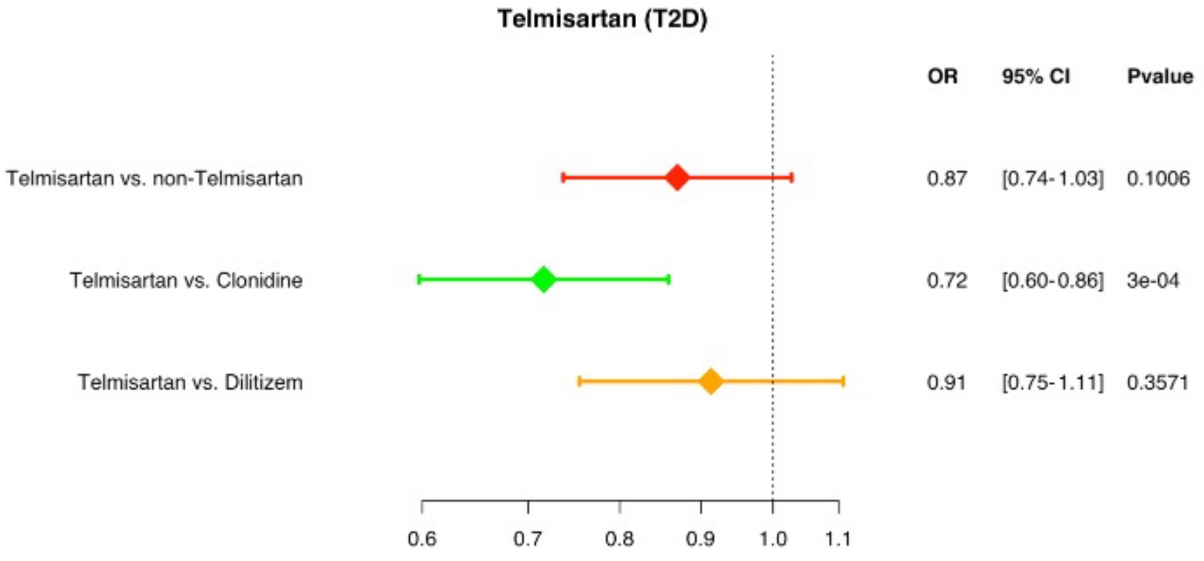


**b)**

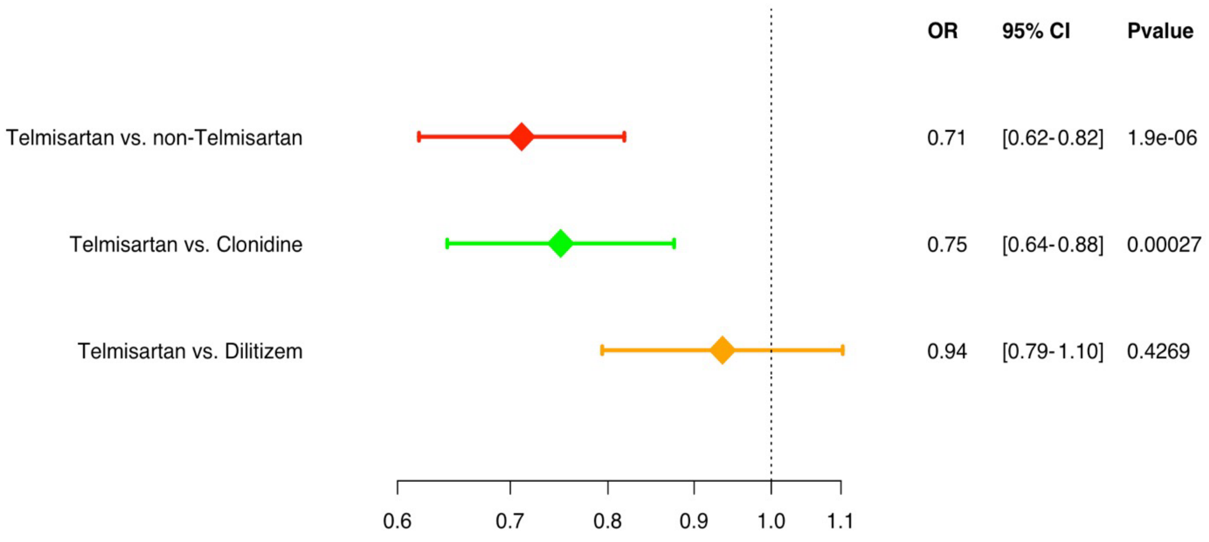


**Supplementary Fig. 2: Sex-specific Hazard ratios (HR)** and 95% confidence interval (CI) across three cohort studies for **(a)** males and **(b)** females**.**

**a)**

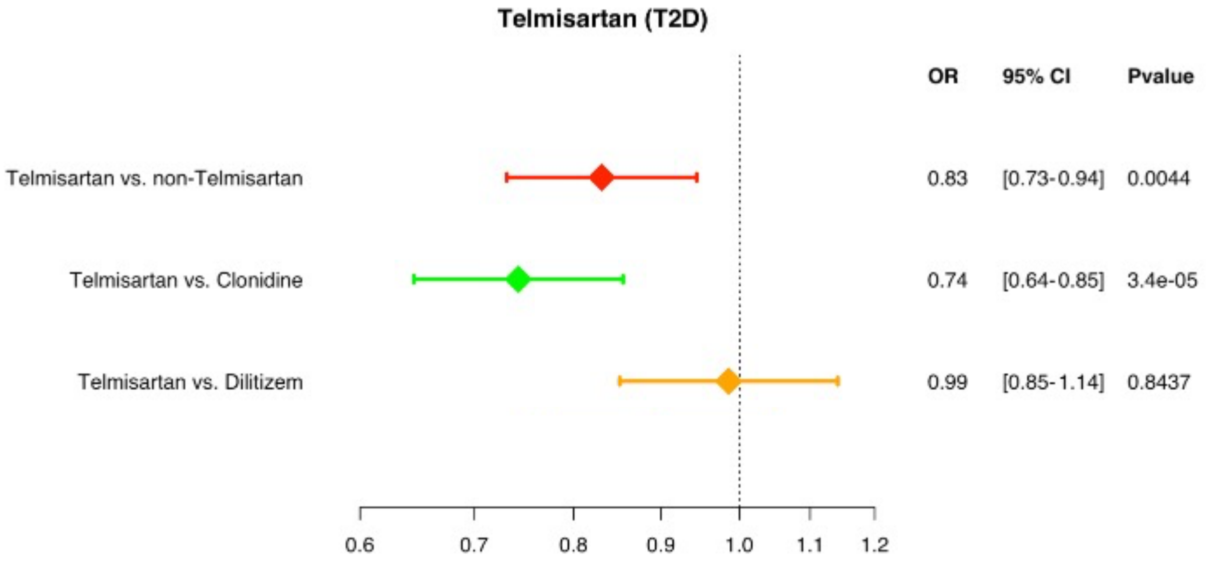


**b)**

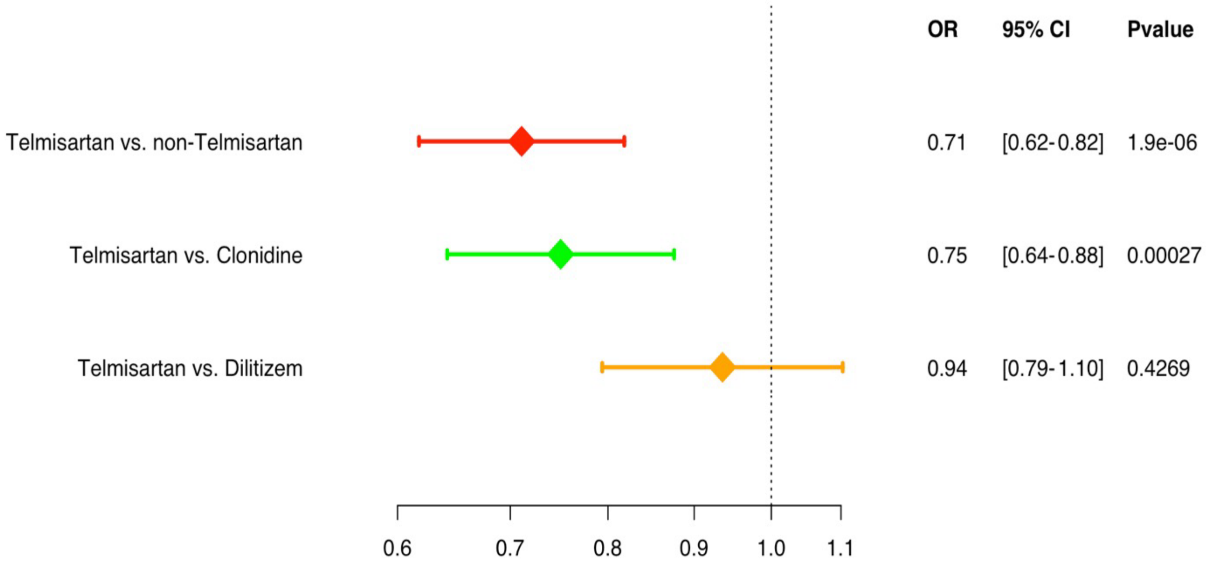


**Supplementary Fig. 3. Age-stratified Hazard ratios (HR)** and 95% confidence interval (CI) across three cohort studies for **(a)** patients >55 years old and **(b)** younger patients of age 20-55 years old.
